# Modification of clinical indicators for differentiating stages of chronic HBV infection based on pathological changes in liver tissue

**DOI:** 10.1101/2023.03.27.23287717

**Authors:** Wentao Li, Binhao Zhang, Jiansheng Zhu, Junyan Liu, Qiupeng Wang, Jiang Feng, Tongjing Xing

## Abstract

**Objective:** To screen and modify more accurate clinical and viral indicators for differentiating the different stages of chronic hepatitis B virus (HBV) infection based on liver histopathological changes.

**Methods:** The clinical and liver pathology data of chronic hepatitis B (CHB) patients undergoing liver biopsy were collected for retrospective analysis. The area under the curve (AUC) of the receiver operating characteristic (ROC) was used to evaluate the diagnostic value for differentiating the different stages of chronic HBV infection.

**Results:** A total of 118 patients who met the diagnostic and exclusion criteria were selected. There were significant differences among HBV DNA, hepatitis B surface antigen (HBsAg), HBeAg, HBcAb, and platelets (PLT) between the IT and IC stages. Platelets were significantly higher in patients with the IT stage of CHB than in patients with the IC stage, whereas HBcAb levels were directly reversed. Multivariate analysis showed that HBeAg independently correlated with the IT and IC stages. Univariate analysis showed that HBV DNA and HBsAg were quantified between the ICO and IR stages except for ALT. The cutoff value of HBeAg used to quantitatively differentiate between IT and IC was 1335 and the AUC was 0.921 (95% confidence interval (CI): 0.836 to 0.971).

**Conclusions:** The high levels of HBeAg rather than HBeAg positive might help to identify patients with the “true” IT stage. PLT and HBcAb are effective indicators for differentiating patients between the IT and IC stages of chronic HBV infection. HBV DNA of <20 IU/mL may be a more rational cutoff value for ICO patients.

## Introduction

According to a report from the World Health Organization, 20–30% of the approximately 240 million chronic hepatitis B virus (HBV) infections worldwide develop into cirrhosis and/or hepatocellular carcinoma (HCC) [1]. In the course of chronic HBV infection, the long-term interaction between the host and the HBV leads to the repeated fluctuation of intrahepatic inflammation and the occurrence of liver fibrosis and disease progression [2]. Based on the changes in alanine aminotransferase (ALT), HBV DNA, hepatitis B e-antigen (HBeAg), and the liver pathological damage, the natural history of chronic HBV infection is divided into four stages, namely immune tolerance (IT), immune clearance (IC), immune control (ICO), and immune reactivity (IR). There are different virological, biochemical, and histological characteristics at the four stages. This classification method plays an important role in guiding the diagnosis, treatment, and prognosis evaluation of chronic HBV infection [3, 4].

Studies have shown that the HBV-specific immune response determines the outcome of HBV infection [5]. Dysregulated or exhausted virus-specific T and B cell functions persist during chronic HBV infections, but no distinct immune signature based on T and B cells is available for clinical subtyping [6]. Recently, Park et al. [7] found the presence of HBV-specific T cells in patients with the IT stage. Mason et al. [8] found the presence of hepatocyte clones within the liver tissue of patients in the IT stage. A large number of liver biopsy studies have found different degrees of inflammation and fibrosis in some patients with the so-called “immune tolerance.” Some scholars have raised different opinions on the IT stage of chronic HBV infection [11]. In view of this, in 2017, the European Society for Liver Disease (EASL) proposed a reclassification of the natural history of chronic HBV infection into five stages instead of the previous four stages [12]. However, there is no international consensus on this new classification [13, 14].

The guidelines issued by the EASL, American Association for the Study of Liver Diseases, Asian Pacific Association for the Study of the Liver, and the Chinese Society of Liver Diseases and Infectious Diseases Society made recommendations for the staging of the natural history of chronic HBV infection. The main evaluation indicators used are consistent, such as ALT, HBV DNA, HBeAg, and hepatitis B surface antigen (HBsAg) quantification. However, there are still obvious differences in the definition level of each indicator. In particular, HBeAg positive and negative and not quantitative detection were used to differentiate the four stages [12, 15, 16, 17]. These differences led to the inconsistency in the inclusion criteria of patients in many basic and clinical studies and affected the evaluation of the diagnosis and prognosis of patients with chronic hepatitis B (CHB) [18, 19, 20]. Further unification needs to be explored. Therefore, based on the histopathological changes in the liver, the level of clinical and viral indicators between the IT and IC stages or ICO and IR stages were analyzed and modified to provide more reasonable guidance for the staging and treatment of CHB.

## Patients and Methods

### Patients

The clinical and pathological data of patients with CHB who underwent liver biopsies were collected at Taizhou Hospital of Zhejiang Province affiliated to Wenzhou Medical University from 2015 to 2022. The diagnosis and staging of CHB were performed according to the Chinese Guidelines for the Prevention and Treatment of Chronic Hepatitis B (2019 edition)[15]. The exclusion criteria are as follows: ① patients with hepatitis A, C, D, or E virus infection, cytomegalovirus, Epstein–Barr virus infection, or human immunodeficiency virus infection; ② patients treated with interferon-α or nucleos(t)ide analogs in the last 6 months; ③ patients with moderate to severe fatty liver, alcoholic liver, drug liver, autoimmune, or hereditary liver disease; ④ patients with liver cancer and cancer of other organs; ⑤ patients with decompensated cirrhosis with severe heart, lung, or kidney disease, or diabetes. The staging of CHB was conducted based on their HBsAg, HBV DNA, HBeAg, ALT and liver pathology. The selected patients were categorized into the IT, IC, ICO, and IR stages. The criterion of IT stages as following: HBsAg>1× 10^4^IU/ml, HBeAg positive, HBV DNA>2×10^7^IU/ml, ALT normal, liver pathology without obvious inflammation and fibrosis. The criterion of IC stage as following: HBsAg positive, HBeAg positive, HBV DNA>2×10^4^IU/ml, ALT continuously or repeatedly increased, obvious inflammatory necrosis and/or fibrosis of liver histology. The criterion of ICO stage as following: HBsAg< 1×10^3^IU/ml, HBeAg negative, HBV DNA<2×10^3^IU/ml, ALT normal, no or only mild inflammation and/or different fibrosis of liver histology. The criterion of IR stage as following: HBsAg positive, HBeAg negative, HBVDNA ≥2×10^3^IU/ml, ALT continuously or repeatedly increased and obvious inflammatory necrosis and/or fibrosis of liver histology[15]. This study was approved by the Ethical Commission of Taizhou Hospital of Zhejiang Province.

### Liver biopsy and pathological diagnosis

Liver samples were obtained using a 16-gauge Menghini needle that was ultrasound-guided. The length of the biopsy specimen was longer than 15 mm. Hematoxylin and eosin (H&E) staining and Masson staining were routinely performed. The inflammation grade and fibrosis stage of the liver biopsy specimens were diagnosed according to the Scheuer scoring system[15]. The degree of liver inflammatory necrosis was categorized into mild (0–G1), moderate (G2–G3), and severe (G4). The degree of liver fibrosis was categorized as mild (0–F1), moderate (F2–F3), and severe (F4). All patients signed an informed consent form.

### Detection of serum markers and HBV DNA quantification

Serum HBsAg levels were quantified using an automated chemiluminescence immunoanalyzer. The kit was purchased from Abbott Laboratories. The range of detection was 0.05–250 IU/mL. If the sample concentration was more than 250IU/mL, the sample was diluted and retested. Semiquantitative detection of serum HBeAg and hepatitis B core antibody (HBcAb) (S/co) levels was performed using an automated chemiluminescence immunoanalyzer. Serum HBV DNA was detected using a quantitative PCR instrument (ABI, USA). The kit was purchased from Shanghai Haoyuan Biological Technology Co., Ltd. The lower detected limit was 20 IU/mL.

### Statistical analysis

All statistical analyses were performed using SPSS 26.0 (IBM, NY, USA). Data meeting the normal distribution were expressed as mean ± standard deviation. Student’s *t*-test was used to compare the IT versus IC periods and ICO versus IR periods. Data not normally distributed are shown as median (quartile interval). The Mann–Whitney *U* test was used to test the IT and IC periods and the ICO and IR stages. For categorical variables, the number of cases (percentage) was indicated and the chi-squared test or Fisher’s exact test was performed. The Hosmer–Lemeshow test was used to assess the goodness of fit of the model. The area under the curve (AUC) of the receiver operating characteristic (ROC) was used to evaluate the efficacy of the model during the IT and IC and ICO and IR stages with a preset cutoff using the maximum Youden index. Differences between the ROC were compared using the Deron test. Significance was set at P < 0.05.

## Results

### Baseline data for the collected patients

The data of 118 patients, comprising 83 males and 35 females, who met the diagnostic and exclusion criteria were collected and used in this analysis. The mean age is 43 years, and 76 HBeAg-positive and 42 HBeAg-negative patients were selected. The baseline clinical data and pathological changes in liver tissue are shown in Table 1. The staging was categorized based on the clinical and pathological criteria of guidelines.

**Table 1.** Baseline data for collected patients.

| Variable |  | n=118 |
| --- | --- | --- |
| Sex ‡ | Male/Female | 83 (70.3)/35 (29.7) |
| Age * |  | 43 ( 11 ) |
| ALT(U/L) # |  | 53.50 [33.50, 75.25] |
| AST(U/L) # |  | 43.00 [29.00, 59.00] |
| Albumin(g/L)* |  | 41.65 (4.10) |
| Platelet (×10<sup>9</sup>/L) # |  | 174.00 [133.00, 207.00] |
| Total bilirubin(μmol/L) # |  | 14.10 [11.2, 18.95] |
| HBVDNA(LogIU/ml) # |  | 7.06 [5.55, 7.94] |
| HBsAg(LogIU/ml) # |  | 3.52 [2.98, 4.32] |
| HBeAg(s/co) # |  | 81.63 [0.43, 1364.44] |
| HBcAb(s/co) # |  | 8.91 [7.53, 9.90] |
| Inflammation grade‡ | G1 | 36 (30.5) |
|  | G2 | 79 (67) |
|  | G3 | 3 ( 2.5) |
| Fibrosis stage‡ | F0 | 1 ( 0.9) |
|  | F1 | 81 (68.6) |
|  | F2 | 35 (29.6) |
|  | F3 | 1 ( 0.9) |

**Legend:** \*Mean ± standard deviation; ‡ Percentage; # Median and interquartile range

### Comparative analysis of different stages of chronic HBV infections

Twenty-one patients with the IT stage and 55 patients with the IC stage constitute the 76 HBeAg-positive patients. Fourteen patients with the ICO stage and 28 patients with the IR stage constitute the 42 HBeAg-negative patients. Univariate analysis shows a significant difference among HBV DNA, HBsAg, HBeAg, HBcAb, and platelets (PLT) between the IT and IC phases except for ALT. There is no significant difference among age, sex, albumin, and total bilirubin between the two groups. Multivariate analysis shows that HBeAg independently correlate with the stages(Table 2). The PLT levels in patients with the IT stage are higher than those in patients with the IC stage, 205.00 [177.00, 238.00] × 10^9^/L and 165.00 [134.00, 207.00] × 10^9^/L, respectively. The semiquantitative levels of HBeAg are significantly higher in patients with the IT phase than in patients with the IC phase, 1588.93 [1433.06, 1799.06] s/co and 740.55 [53.01, 1169.98] s/co, respectively. The semiquantitative level of HBcAb is 6.88 [4.83, 8.88] s/co in the IT stage and 9.11 [7.94, 9.79] s/co in the IC stage (P < 0.001) (Table 3). Univariate analysis shows a significant difference among HBV DNA and HBsAg between the ICO and IR stages except for ALT. There is no significant difference among age, sex, albumin, total bilirubin, PLT, and HBcAb between the two groups (Table 4).

**Table 2.** Comparative analysis of IT stage and IC stage of patients with CHB.

| Variables |  | Overall(n=76) | IT stage (n=21) | IC stage (n=55) | P |
| --- | --- | --- | --- | --- | --- |
| Sex‡ | Male | 49 (64.5) | 11 ( 52.4) | 38 (69.1) | 0.173 |
|  | Female | 27 (35.5) | 10 ( 47.6) | 17 (30.9) |  |
| Age * |  | 41.61 ( 11.44) | 38.67(7.38) | 42.72(12.53) | 0.168 |
| ALT(U/L) # |  | 57.00[36.25,75.00] | 26.00[19.50, 32.00] | 69.00[55.00, 93.00] | <<br>0.001 |
| AST(U/L) # |  | 43.00 [31.5, 59.75] | 25.00 [22.5,28.00] | 52.00 [41.00, 78.00] | <<br>0.001 |
| Albumin(g/L) * |  | 41.20 (4.28) | 42.64(3.73) | 40.65 (4.37) | 0.069 |
| Platelet (×10<sup>9</sup>/L) # |  | 181.50[145.00, 211.50] | 205.00[177.00,238.00] | 165.00[134.00,207.00] | 0.003 |
| Total bilirubin(μmol/L) # |  | 13.9[10.75,18.78] | 13.30 [10.65, 16.65] | 15.10 [10.9, 18.90] | 0.267 |
| HBVDNA (LogIU/ml) # |  | 7.76[6.98,8.25] | 8.28[7.83,8.51] | 7.35[6.27,7.96] | <<br>0.001 |
| HBsAg (LogIU/ml) # |  | 3.97[3.23,4.61] | 4.79[4.50,4.91] | 3.49[3.05,4.27] | <<br>0.001 |
| HBeAg(s/co) # |  | 1061.96[104.13,1490.79] | 1588.93[1433.06,1799.06] | 740.55[53.01,1169.98] | <<br>0.001 |
| HBcAb(s/co) # |  | 8.81 [7.12,9.57] | 6.88 [4.83,8.88] | 9.11[7.94,9.79] | <<br>0.001 |
| Inflammation grade ‡ | G1 | 22 (29) | 21 (100) | 1 (1.8) | <<br>0.001 |
|  | G2 | 52 (68.4) | 0 ( 0.0) | 52 (94.6) |  |
|  | G3 | 2(2.6) | 0 ( 0.0) | 2 (3.0) |  |
| Fibrosis stage‡ | F1 | 55 (72.4) | 21 (100.0) | 34 (61.8) | 0.004 |
|  | F2 | 20 (26.3) | 0 ( 0.0) | 20 (36.4) |  |
|  | F3 | 1 (1.3) | 0 ( 0.0) | 1 ( 1.8) |  |

**Legend:** \*Mean ± standard deviation; ‡ Percentage; # Median and interquartile range

**Table 3.**
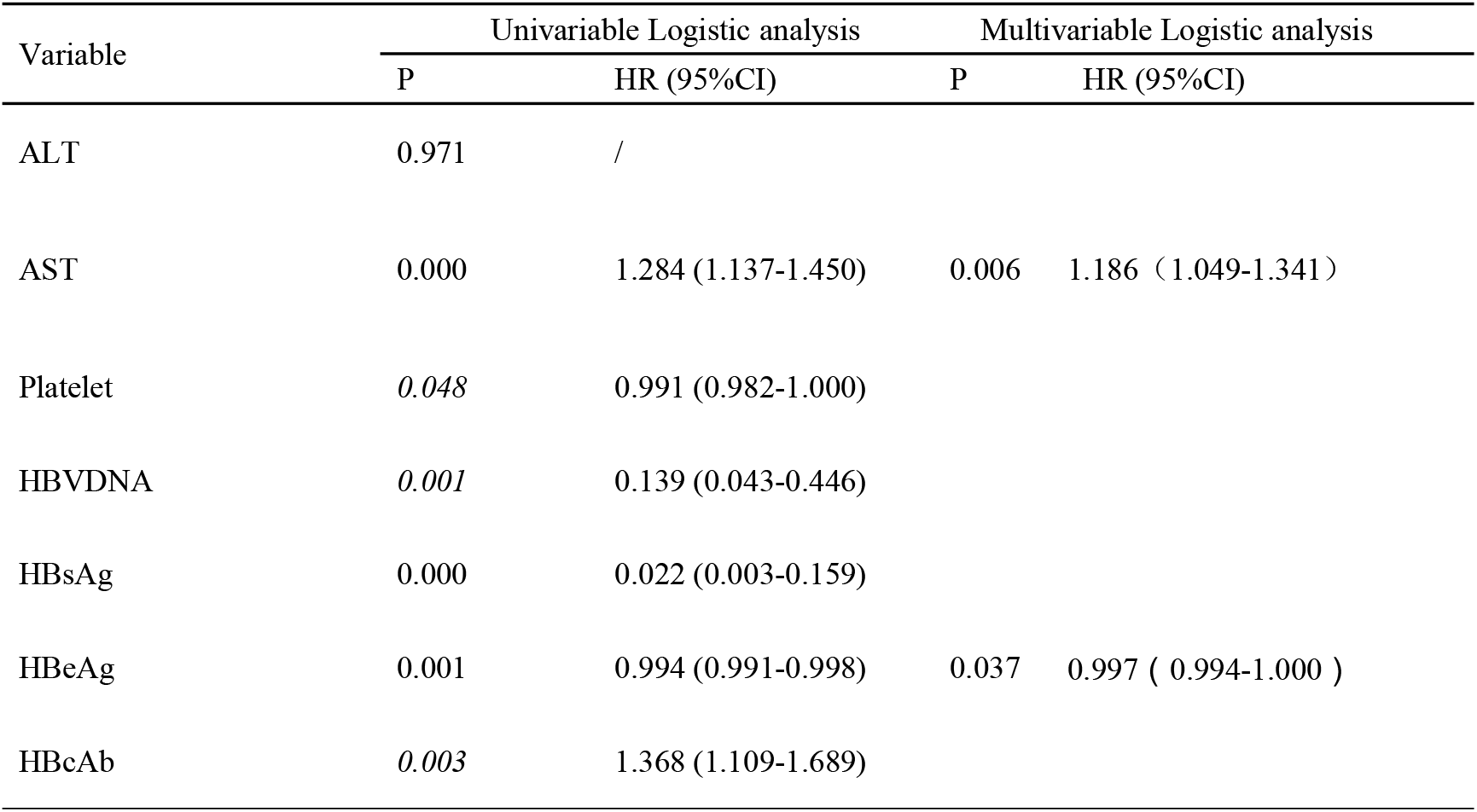
Univariate and multivariate analyses in factors related with IT and IC stage of CHB patients.

**Table 4.** Comparative analysis of ICO stage and IR stage of patients with CHB.

| Variable |  | Overall (n=42) | ICO(n=14) | IR (n=28) | P |
| --- | --- | --- | --- | --- | --- |
| Sex<sup>‡</sup> | Male | 34 (81) | 12 ( 85.7) | 22 (78.6) | 0.890 |
|  | Female | 8 (19) | 2 ( 14.3) | 6 (21.4) |  |
| Age * |  | 44.24 ( 8.84) | 46.50 ( 8.74) | 43.11 ( 8.82) | 0.246 |
| ALT(U/L) # |  | 49.00 [24.75, 77.25] | 20.00 [15.75, 25.25] | 61.00 [48.5, 109.25] | <0.001 |
| AST(U/L) # |  | 40.50 [24.75, 52.25] | 23.00 [21.50, 26.25] | 50.00 [39.25, 76.75] | <0.001 |
| Albumin(g/L)* |  | 42.46 (3.67) | 42.73 (3.70) | 42.33 (3.71) | 0.743 |
| Platelet (×10<sup>9</sup>/L) # |  | 153.5 (50.41) | 173.71 ( 55.23) | 143.39 ( 45.52) | 0.065 |
| Total bilirubin(μmol/L) # |  | 14.4 [11.8, 21.1] | 12.95 [10.58, 17.63] | 15.80 [11.80, 22.40] | 0.196 |
| HBVDNA (LogIU/ml) # |  | 4.84 [2.72, 6.61] | 2.03 [1.30, 2.84] | 5.84 [4.76, 6.83] | <0.001 |
| HBsAg (LogIU/ml) # |  | 3.04 [2.34, 3.54] | 1.96 [0.63, 2.84] | 3.31 [3.03, 3.56] | <0.001 |
| HBcAb(s/co) # |  | 9.35 [8.29, 10.49] | 9.73 [8.29, 10.59] | 9.15 [8.14, 10.34] | 0.408 |
| Inflammation grade<sup>‡</sup> | G1 | 14 (33.3) | 14 (100.0) | 0 ( 0.0) |  |
|  | G2 | 27 (64.3) | 0 ( 0.0) | 27 (96.4) |  |
|  | G3 | 1 ( 2.4) | 0 ( 0.0) | 1 ( 3.6) |  |
| Fibrosis stage<sup>‡</sup> | F0 | 1 ( 2.4) | 1 ( 7.14) | 0 ( 0.0) |  |
|  | F1 | 26 (61.9) | 10 ( 71.43) | 16 (57.1) |  |
|  | F2 | 15 (35.7) | 3 ( 21.43) | 12 (42.9) |  |

**Legend:** \*Mean ± standard deviation; <sup>‡</sup> Percentage; <sup>#</sup> Median and interquartile range

### ROC curve analysis in the IT and IC stages of CHB patients

ROC curve analysis of the indicators of the IT and IC stages was performed using Medcalc software. The AUC is 0.806 (95% confidence interval (CI): 0.699–0.888) using HBV DNA as a variable. The cutoff value under the maximum Yoden index is 7.509 for LOG HBV DNA. The sensitivity and specificity are 56.36% and 100.00%, respectively (Figure 1A). The HBV DNA cutoff value was used to differentiate 45 patients with the IT stage and 31 patients with the IC stage. The cutoff value of the HBsAg quantitative is more than 4.006. The AUC is 0.912 (95% CI: 0.824–0.965). The sensitivity and specificity are 70.91% and 100.00%, respectively (Figure 1B). Thirty-eight patients with the IT stage and 38 patients with the IC stage were differentiated based on the cutoff value of HBsAg. The cutoff value of the HBeAg quantitative is 1335.57. The AUC is 0.921 (95% CI: 0.836–0.971). The sensitivity and specificity are 80.0% and 100%, respectively (Figure 1C). Thirty-two patients with the IT stage and 44 patients with the IC stage were differentiated based on the cutoff value of HBeAg. HBcAb was used as a variable to differentiate the IT stage from the IC stage, yielding an AUC of 0.778 (95% CI: 0.665–0.868) with a maximum cutoff of 7.1 (S/co) (Figure 1D). Nineteen patients with the IT stage and 57 patients with the IC stage were differentiated based on the cutoff value of HBcAb. The cutoff value under the maximum Jordan index is 161 × 10^9^/L. The AUC of the PLT is 0.720 (95% CI: 0.605–0.817). The sensitivity and specificity are 49.09% and 100.00%, respectively (Figure 1E). There are 28 patients with the IT stage and 48 patients with the IC stage were differentiated based on the cutoff value of the PLT.

**Figure 1.**
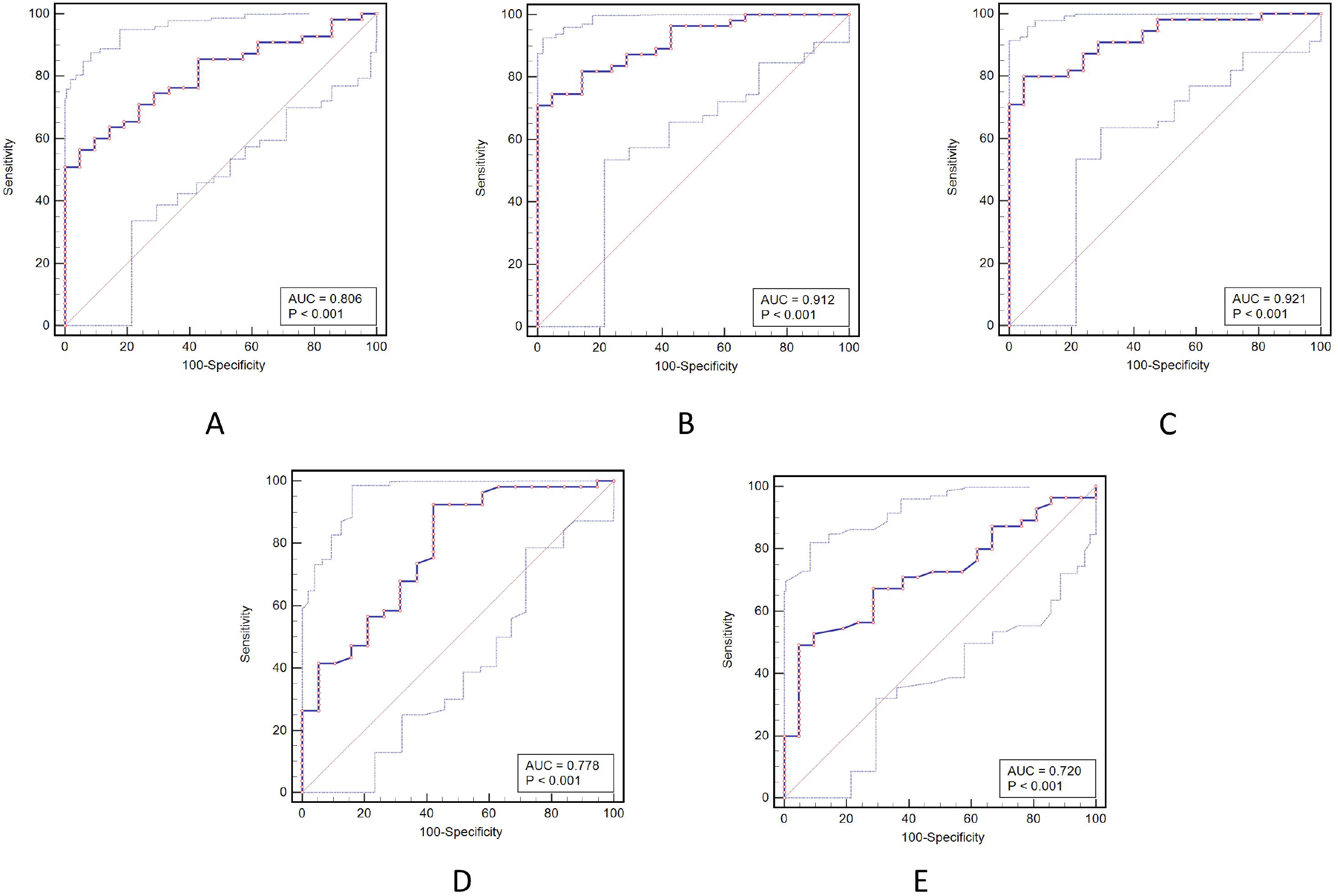
ROC curve analysis in the IT and IC stages of CHB patients. A, HBV DNA; B, HBsAg; C, HBeAg; D, HBcAb; E, PLT

### ROC curve analysis of patients during the ICO and IR stages

Analysis of the ROC curve was performed using the Medcalc software. LOG HBV DNA was used as a variable to differentiate the ICO and IR stages. The cutoff value of the LOG HBV DNA under the maximum Youden index is more than 3.279. The AUC of the ROC curve is 1.000 (95% CI: 0.916–1.000). The sensitivity and specificity are 100.00% and 28.57%, respectively (Figure 2A). Fifteen patients with the ICO stage and 28 patients with the IR stage were differentiated using the cutoff value of the HBV DNA. The cutoff value of the LOG HBsAg under the maximum Yoden index is more than 2.934. The AUC of the ROC curve is 0.967 (95% CI: 0.860–0.998). The sensitivity and specificity are 89.29% and 18.12%, respectively (Figure 2B). Sixteen patients with the ICO stage and 26 patients with the IR stage were differentiated based on the LOG HBsAg cutoff value.

**Figure 2.**
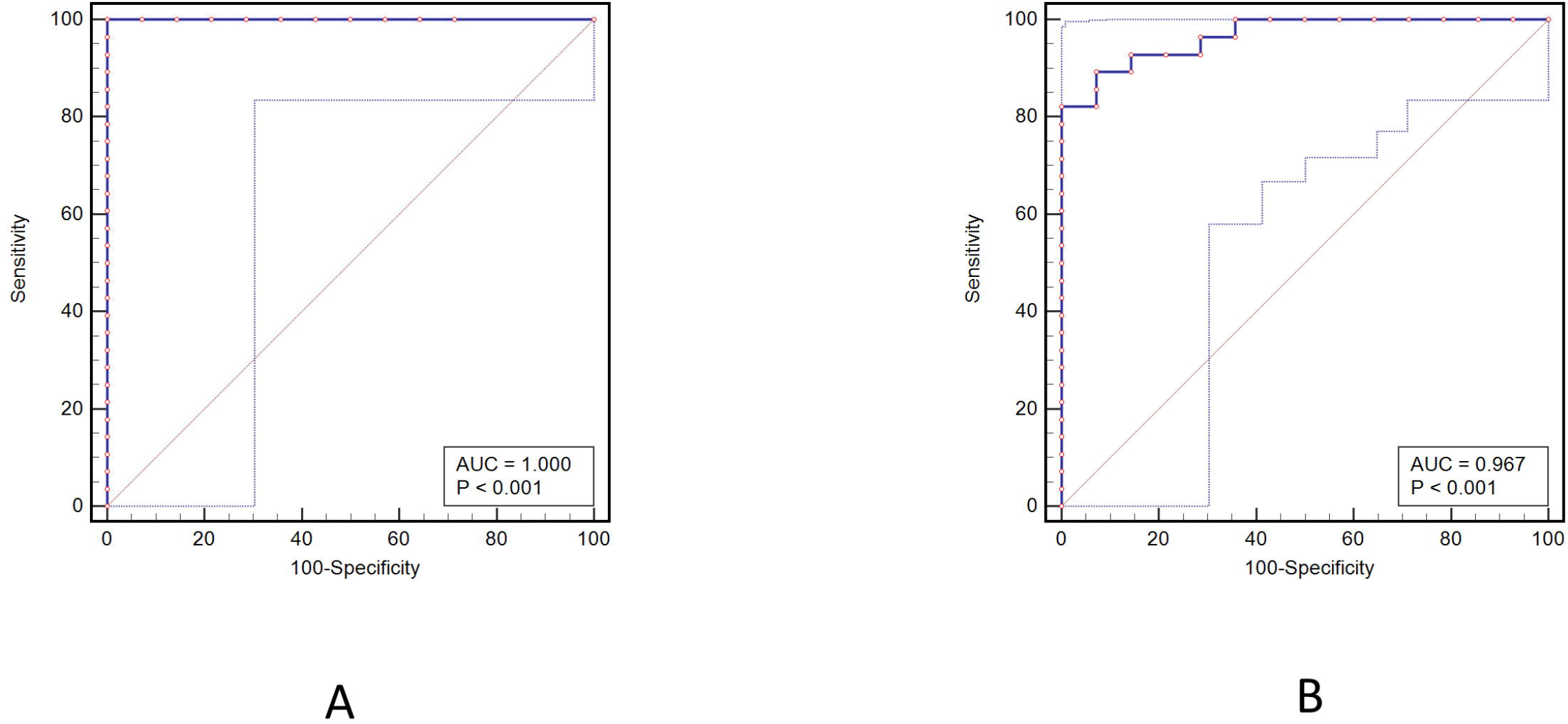
ROC curve analysis in the ICO and IR stages of CHB patients. A, HBV DNA; B, HBsAg

### Comparative analysis of patients with “true” ICO and existing patients with the ICO stage

The level of HBV DNA is less than 2 × 10^3^ IU/mL and that of HBsAg is less than 1 × 10^3^ IU/mL in patients with the ICO stage. The World Gastroenterology Organization (WGO) considers the HBV DNA level of patients with the ICO stage to be less than 200 IU/mL. In this study, the median of the HBV DNA quantitative level in the ICO stage is 1.92 [1.30, 2.71]. The median of the HBsAg quantitative level is 1.83 [0.40, 2.82]. Patients with the ICO stage are divided into two groups based on an HBV DNA of ≥ 20 IU/mL and < 20 IU/mL. The characteristics of the two groups were analyzed (Table 5). The results show that patients with an HBV DNA of < 20 IU/mL have low levels of HBsAg. The grades of inflammation and stages of fibrosis of the liver tissues in 3 of 4 patients are mild (G1F0-1). The liver fibrosis of 1 patient is moderate (F2). Ten patients with an HBV DNA of ≥ 20 IU/mL have relatively high levels of HBsAg. The grades of inflammation of the liver tissues in 10 patients are mild (G1). The stages of fibrosis of the liver tissues in 1, 7, and 2 patients are F0, F1, and F2, respectively.

**Table 5.**
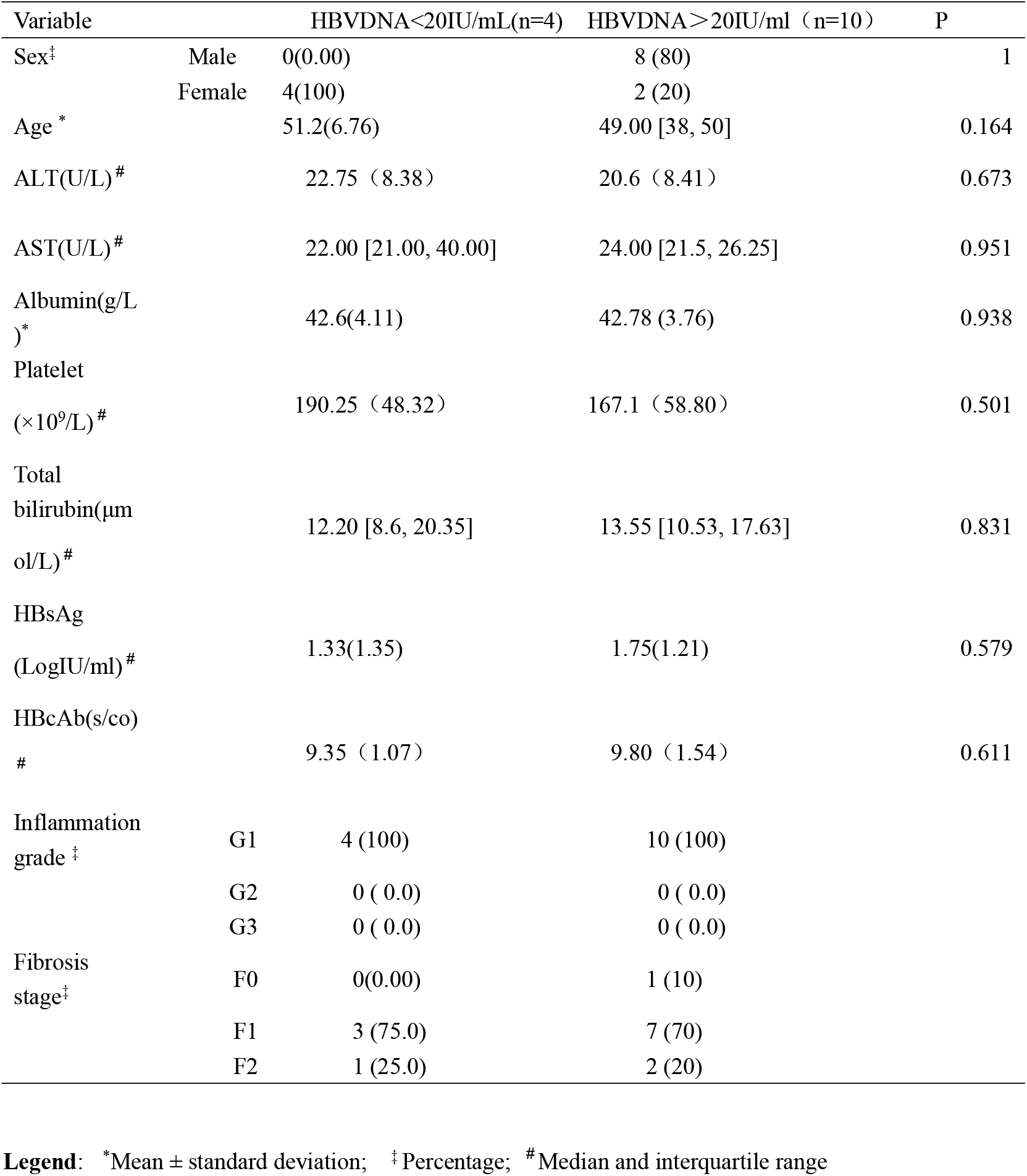
Comparative analysis of ICO stage of CHB patients between HBVDNA<20 and ≥ 20 IU/ml.

## Discussion

At present, the indicators used for staging chronic HBV infection mainly comprise ALT, HBV DNA, HBeAg, HBsAg, and liver pathological changes. However, owing to the invasiveness of liver biopsy, most of the related studies did not include the liver pathological status, which affected the accuracy of the clinical staging of CHB patients to a large extent [4,15]. This study analyzed the indicators used in the clinical staging of chronic HBV-infected patients based on the liver histopathological changes in patients with chronic HBV infection. The biochemical and virological indicators commonly used in patients are different in various stages of CHB patients. The high level of HBeAg semiquantification is especially important to the definition of the IT stage. The levels of HBcAb and PLT are significantly different in the IT and IC stages and could be used as indicators to differentiate the IT and IC stages.

The large gap in the definition of the IT and IC stages has influenced the comparison of the results of different studies and the evaluation of the effect of anti-HBV therapy [4,13]. In this study, the results show that there are significant differences in the levels of ALT, HBV DNA, HBsAg, HBeAg, PLT, and HBcAb between the IT and IC stages. Considering HBeAg is a marker of the initiation of the immune response in patients with CHB, a decline in HBeAg levels suggests that CHB patients have entered the IC phase [25]. The fact of being HBeAg-positive (i.e., S/co > 1), and not the difference in HBeAg levels as a differentiation criterion of the IT and IC stages, might result in the problem of inaccurate staging. Based on the pathological changes in the liver, this study found the cutoff value of the semiquantitative level of HBeAg that differentiated the IT and IC periods to be 1341.36 S/co. The IT stage differentiated based on the HBeAg cutoff value of this study might be more in line with the definition of the IT stage and more helpful to guide the treatment of CHB.

The standards of HBV DNA established by the various guidelines are also inconsistent, ranging from 1 × 10^6^ IU/mL to > 2 × 10^7^ IU/mL. The standard identified by the WGO is more than 1 × 10^9^ IU/mL [26]. HBsAg quantification at the IT stage level is mostly more than 1 × 10^4^ IU/mL. These relevant criteria lack evidence-based medical evidence of a large sample and whether it reflects the “immune tolerance” state. Hui et al. reported that patients with liver fibrosis of more than F1 were excluded from the IT stage. Patients who remained in the IT stage only presented with mild lesions during the follow-up period. The median serum HBV DNA is 9.81 log copies/mL in 48 immune-tolerant patients. The results of this study show that the cutoff value of HBV DNA is > 7.51 log IU/mL and similar to the results reported by Hui et al. [27]. The EASL guidelines suggest that the level of HBsAg quantification should be high, but the specific values are not supported. Tseng et al. suggested that the level of HBsAg should be more than 1 × 10^5^ IU/mL [28]. This study found that the cutoff value for HBsAg quantification in patients with the IT stage is > 4.01 log IU/mL, which is similar to the criteria of the current domestic guidelines.

Studies have shown that the number of PLT gradually decreased in patients with liver fibrosis and negatively correlated with the degree of liver fibrosis. The decline in PLT level was more obvious, especially in the stage of cirrhosis [29]. Aiolfi et al. [30] showed that PLT plays a key role in the pathogenesis of acute and chronic liver diseases associated with HBV infection by promoting the accumulation of virus-specific CD8^+^T cells and nonspecific inflammatory cells in the liver parenchyma. PLT can activate hepatic stellate cells through the release of 5-serotonin, transforming growth factor-β1, platelet-derived growth factor, and other substances, promoting the formation of liver fibrosis in chronic liver injury [31]. The role of anti-inflammatory agents, such as aspirin, was found in both animal models and clinical studies [32,33]. PLT were an independently associated factor for the staging of the IT and IC phases in chronic HBV-infected patients. The PLT level of patients with CHB IC is significantly lower than that of patients with IT. Patients with the IT and IC stages were differentiated using the cutoff value of the PLT as 161 × 10^9^/L. These results suggest that PLT can be used as one of the indicators to differentiate between patients with the IT and IC stages. The reason may be related to the presence of obvious liver fibrosis in patients with the IC stage, and PLT may play an important role in the development of liver fibrosis and clinical evaluation.

HBcAb is the earliest antibody produced after patients are infected with HBV, which is an important marker of hepatitis B infection. However, it is not a protective antibody [34]. Song et al. [35] found that the quantitative serum levels of HBcAb in chronic HBV-infected patients were closely correlated with hepatitis activity. Studies have shown that the baseline HBcAb levels predict treatment response in HBeAg-positive CHB cohorts [36,37,38].

Jia et al. [39] reported that the levels of HBcAb significantly varied at different stages of chronic HBV patients. The median HBcAb in patients with IT is significantly lower than that in patients with IC. The median HBcAb in the ICO stage is significantly lower than that in the IR stage. Su et al. suggested that HBcAb quantification combined with other indicators could differentiate between patients with the IC and IR stages [40]. Chi et al. [41] showed that the level of HBcAb was associated with relapse after withdrawal of Nucleotide analogues(NAs) treatment. The higher the level of HBcAb, the lower the relapse rate after withdrawal of NAs treatment. This study found that the semiquantitative level of HBcAb in patients with the IT stage was lower than that of patients with the IC stage and similar to the results of Jia et al. [39]. No significant difference was found between the ICO and IR stages, which is inconsistent with the results of Jia et al. [39]. The reasons for the inconsistency might be related to the detection methods and sample differences. Further studies are needed.

The levels of HBsAg and HBV DNA are different in the current criteria for differentiating between patients with the ICO and IR stages [12,42]. Ungtrakul et al. [43] showed that the ICO status of CHB and HBeAg-negative CHB were better differentiated using an HBsAg lower than 1000 IU/mL and an HBV DNA less than 2000 IU/mL. The sensitivity and specificity were 41% and 72%, respectively. This criterion was used to differentiate the ICO patients from the IR patients by most of the current guidelines. However, recent studies have found that patients with HBV DNA levels less than 2000 IU/mL still have a high incidence of cirrhosis and HCC and these patients may also need therapy [44,45]. The results of this study show that the HBV DNA and HBsAg quantitative levels are significantly different between the ICO and IR stages except for ALT. The cutoff value for the quantification of log HBsAg used to differentiate patients with the two stages is 2.93 log IU/mL, which is similar to the HBsAg of < 1000 IU/mL in the CHB guidelines. The cutoff value of HBV DNA is 3.28 log IU/mL, which is also similar to the HBV DNA of < 2000 IU/mL in the CHB guidelines.

The persistent negative HBV DNA and HBsAg are the evaluation criteria for clinical cure. However, many patients still have high levels of HBsAg after negative HBV DNA due to the influence of HBV DNA integration. The 2019 International Conference on Liver Diseases stated that persistently negative HBV DNA, whether HBsAg-positive or not could be used as an intermediate indicator of NAs treatment. In this study, patients in the ICO stage were divided into two groups based on an HBVDNA of < 20 IU/mL and 20 ≥ IU/mL. The results show that patients with an HBV DNA of < 20 IU/mL have low levels of HBsAg. The liver pathology is mild. However, all the patients with an HBV DNA of ≥ 20 IU/mL have higher levels of HBsAg. Most of them have severe liver fibrosis. Some patients may not have obvious tissue damage after the virus clearance considering the mechanisms of HBV clearance may be cytotoxic and noncytotoxic. [46,47]. Therefore, the liver pathological changes assessment for patients with the ICO stage also has some limitations and needs to be combined with the clinical indicators for a comprehensive evaluation.

This study has some limitations. First, this study is a retrospective analysis from a single center. The small sample size makes it difficult to adequately reflect all patients, and further sample expansion and multicenter study validation are needed. Second, liver pathological specimens are accompanied with mild fatty changes in part of the samples, which may affect the accuracy of this study. The detection methods of HBeAg and HBcAb used in this study are semiquantitative, which requires validation using quantitative detection methods. Finally, a dynamic observational analysis of patient prognosis was not performed in this study. This requires further follow-up observation in future studies.

In conclusion, this study optimized the levels of HBsAg, HBV DNA, and HBeAg based on liver pathological changes and proposed new cutoff values for the staging of chronic HBV infection, especially the HBeAg semiquantitative is greater than 1335 S/co, which can help to clinically differentiate between patients with the “true” IT stage. PLT and HBcAb are found to be good indicators for differentiating patients with the IT and IC stages of chronic HBV infection. HBV DNA of < 20 IU/mL may be the cutoff value for differentiating CHB patients with “true” ICO. These results might provide more accurate criteria for the clinical staging and evaluation of the treatment response of patients with CHB.

## Data Availability

All data produced in the present work are contained in the manuscript.

## Acknowledgments

We thank LetPub (www.letpub.com) for its linguistic assistance during the preparation of this manuscript.

